# Spatial epidemiology of Japanese encephalitis virus and other infections of the central nervous system in Lao PDR (2003 – 2011): a retrospective analysis

**DOI:** 10.1101/19005884

**Authors:** Sayaphet Rattanavong, Audrey Dubot-Pérès, Mayfong Mayxay, Manivanh Vongsouvath, Sue J Lee, Julien Cappelle, Paul N. Newton, Daniel M. Parker

**Author notes:** corresponding author Daniel M. Parker. Sayaphet Rattanavong, Audrey Dubot-Pérès, Mayfong Mayxay, Manivanh Vongsouvath, Sue J Lee, Julien Cappelle, Paul N. Newton.

## Abstract

**Background:** Central nervous system (CNS) infections are important contributors to morbidity and mortality and the causative agents for ∼50% patients are never identified. The causative agents of some CNS infections have distinct spatial and temporal patterns.

**Methodology/Principal Findings:** Here we present the results of a spatial epidemiological and ecological analysis of CNS infections in Lao PDR (2003 – 2011). The data came from hospitalizations for suspected CNS infection at Mahosot Hospital in Vientiane. Out of 1,065 patients, 450 were assigned a confirmed diagnosis. While many communities in Lao PDR are in rural and remote locations, most patients in these data came from villages along major roads. Japanese encephalitis virus ((JEV); n=94) and *Cryptococcus* spp. (n=70) were the most common infections. JEV infections peaked in the rainy season and JEV patients came from villages with higher surface flooding during the same month as admission. JEV infections were spatially dispersed throughout rural areas and were most common in children. *Cryptococcus* spp. infections clustered near Vientiane (an urban area) and among adults.

**Conclusions/Significance:** The spatial and temporal patterns identified in this analysis are related to complex environmental, social, and geographic factors. For example, JEV infected patients came from locations with environmental conditions (surface water) that are suitable to support larger mosquito vector populations. Most patients in these data came from villages that are near major roads; likely the result of geographic and financial access to healthcare and also indicating that CNS diseases are underestimated in the region (especially from more remote areas). As Lao PDR is undergoing major developmental and environmental changes, the space-time distributions of the causative agents of CNS infection will also likely change. There is a major need for increased diagnostic abilities; increased access to healthcare, especially for rural populations; and for increased surveillance throughout the nation.

**AUTHOR SUMMARY:** Infections of the central nervous system (CNS) are important with regard to public health. However many CNS infections are never diagnosed. In this analysis we investigated spatial and temporal patterns in hospitalized patients with suspected CNS infections in Lao PDR. We found that patients were most likely to come from villages located along major roads and highways. Patients from remote areas may have more difficulty reaching healthcare facilities. The most commonly diagnosed infection in these patients was Japanese encephalitis virus (JEV). Patients with this disease came from locations that were optimal for the mosquito vectors that spread JEV, rural areas with surface water and during the rainy season. Our analyses suggest that CNS infections should be a priority for public health workers in the region. Diagnostic capabilities should be increased throughout the nation; surveillance efforts should be broadened; and efforts should be increased toward providing easy access to healthcare for rural and remote populations.

## INTRODUCTION

Numerous illnesses go undiagnosed and the causative agents of many infections are never identified. In regions where access to healthcare facilities is limited and where diagnostic capabilities are few, a smaller proportion of diseases are objectively diagnosed. Diseases with mild symptoms may more frequently go untreated, but in some areas even severe illnesses commonly go undiagnosed and untreated. Infections of the central nervous system (CNS) can be particularly severe, affecting the brain and/or spinal cord and/or the surrounding meninges, frequently resulting in death.

Pathogens that invade and infect the CNS include viruses, bacteria, fungi, parasites, and prions. As with other infections, the causative agent(s) of many CNS infections are never determined (frequently <50%) [1–3]. Symptoms of CNS infection can range widely, even for single causative agents, leading to further difficulties with diagnosis. Diagnoses are frequently presumptive and non-specific (i.e. meningitis is often presumed to be caused by bacteria whereas encephalitis is presumed to be caused viruses [2,4–6]). Some causative agents are specific to regions (e.g. Japanese Encephalitis, Saint Louis Encephalitis, Rift Valley Fever Viruses) and exhibit seasonal fluctuations (e.g. vector borne infections), therefore geography and seasonality can facilitate presumptive diagnosis of CNS diseases [4,7].

In Southeast (SE) Asia, CNS infections are increasingly recognized as important contributors to morbidity and mortality [1,8,9]. However, detailed medical and epidemiological data are frequently lacking, especially for low income nations and from remote areas within middle-to-high income nations. Known important viral CNS infections in SE Asia include Japanese encephalitis, dengue, and rabies viruses [1]. Important bacterial CNS infections include *Streptococcus pneumoniae, Haemophilus influenzae, S. suis, Mycobacterium tuberculosis* and *Neisseria meningitidis. Orientia tsutsugamushi, Rickettsia typhi* and *Leptospira* spp. are increasingly recognized as important causes [9]. Detailed analyses that confirm the cause of CNS related infections or assess their spatial and temporal distribution in the region are rare [8–11,14].

We recently published the results of a study of the etiology and impact of CNS infections diagnosed among 1,065 patients at Mahosot Hospital, Vientiane, Lao PDR [14]. The goal of this secondary analysis was to investigate the spatial distribution(s) of CNS-related infections; to look for differential spatial distributions for major causative agents; and to explore potential geographic, demographic, and environmental correlates of these infections.

## DATA AND METHODS

### Data sources, processing and merging

Data used in this research were compiled from four main sources (**Supporting Figure 1**). The epidemiological data come from an 8-year research project on CNS infections in Lao PDR from all patients who received diagnostic lumbar puncture (LP) at Mahosot Hospital in Vientiane, Lao PDR between January 2003 and August 2011 and consenting to participate [14]. All patients were admitted to the hospital because of suspected CNS infection and Mahosot Hospital is the only medical facility in Lao PDR capable of performing diagnostic LP and cerebral spinal fluid (CSF) analysis. Tests for a large panel of pathogens were performed at the Microbiology Laboratory following international standards (details provided in **Supporting Materials I** and [14]). Demographic (age, gender, ethnicity) and geographic (home village) characteristics of patients were recorded in the database.

The epidemiological data were used to create two separate datasets: One aggregated at the village level (one row per location) and another was maintained at the individual level, with one row per individual. The official Lao PDR censuses from 2005 and 2015 were used to geocode villages (based on village name and administrative units listed in patient records) and to assign village population estimates to each village (taking a mean population size between 2005 and 2015). Village location and population sizes were then merged to both the individual- and village-level datasets.

A subset of villages within the geographic region of the home villages of included patients was selected by overlaying a standard deviational ellipse with 3 standard deviations (calculations described in **Supporting Materials II**) around the patient home villages and then selecting all villages within that ellipse (**Supporting Figure 2**). These villages were then retained for village level comparisons between villages populated, and not populated, with patients admitted with CNS disease needing an LP. This subset of villages is hereafter referred to as the “study area”.

Major road network data was taken from OpenStreetMaps (http://www.openstreetmap.la), selecting “primary”, “secondary”, and all major connecting roads (downloaded in February 2017, **Supporting Figure 3**). Primary and secondary roads are the two largest road classifications for the nation. Primary roads link major towns and cities and secondary roads link mid-sized towns. Primary and secondary link roads are ramps or slip roads that connect other roads to primary or secondary roads. Together, these types of roads are hereafter referred to as “major roads”. Smaller roads were not included in this analysis as they are less likely to be accurately included in the OpenStreetMaps data. The Euclidian distance was then calculated from all villages in the census to the nearest point along a major road. These distances were merged to both the village- and the individual-level datasets.

Environmental predictor variables for vegetation and surface water were derived from Moderate Resolution Imaging Spectroradiometer (MODIS) products (MOD13Q1/MYD13Q1 250 meter AQUA/TERRA 16 day composites). Since many infectious diseases, especially vector borne diseases, are strongly influenced by environmental factors we hypothesized that indicators of vegetation and surface flooding would correlate with some specific diagnoses. Three environmental indices were downloaded and considered in these analyses: a normalized flooding index (NFI) [15]; the normalized difference vegetative index (NDVI); and the enhanced vegetation index (EVI). NFI is indicative of surface water, NDVI is indicative of green surface vegetation, and EVI is an improved measure of green vegetation that is intended to account for dense forest canopies and atmospheric conditions that can lead to error in NDVI measurements. Data were downloaded for each of these environmental indices (EI) within each 16-day time period from February 2002 through December of 2011. The final analyses conducted in this research retained only the EVI and NFI for environmental predictors (summary statistics for NDVI are included) because NDVI and EVI were strongly correlated. The EIs are described in more detail in **Supporting Materials III**.

The environmental raster data were then summarized and extracted based on varying buffer sizes (2km, 5km, and 10km) for each village in the individual- and village-level datasets. Permanent water bodies (such as the Mekong River and Nam Ngum Dam) were masked from the NFI calculations. For the village-level datasets, mean values of each environmental variable was calculated for the study period duration and used as an indicator of “average” vegetation or surface water characteristics of each village. For the individual-level dataset the values were extracted based on the admission date, using increasing durations of time prior to admission (within the same month, within the previous 2 months, within the previous year).

The final datasets include the village-level data, that is a subsample of 98% of all villages with patients included in the study and the other census designated villages within the same region (the study area), and the individual level dataset that includes all patients included in the study. Variables used in this analysis and their descriptions are listed in **Table 1**.

**Table 1:** List of variables, their spatial and temporal scales, and transformations

| Variable | description | spatial scale | temporal scale | transformation |
| --- | --- | --- | --- | --- |
| *Village population* | Population estimate of LP patient home village. This is calculated as a mean population value from the Lao PDR official census from years 2005 and 2015. |  |  | For multivariable regressions, this variable was centered on its mean and standardized by its standard deviation. |
| *Distance to major road* | This is the distance in meters from the LP patient home village and the nearest point on a major highway network, from the OpenStreetMaps map layer. This distance was transformed into kilometers and rounded to the nearest 5 kilometers in order to account for measurement error. | This distance was transformed into kilometers and rounded to the nearest 5 kilometers in order to account for measurement error. |  | For multivariable regressions, this variable was centered on its mean and standardized by its standard deviation. |
| *Village elevation* | This is the elevation of the LP patient home village, calculated from a digital elevation model. It is in meters above sea level. | at village point |  | For multivariable regressions, this variable was centered on its mean and standardized by its standard deviation. |
| *NFI* | This is the normalized flooding index, described in detail in the Supporting materials. | Mean values at 2km, 5km, and 10km buffers around each village. | For village level analysis: calculated as a mean value for the study duration. For individual level analysis: Calculated for the same month (same 16 day time period); the previous 2 months (mean of the previous 5 16 day intervals); and the previous year (mean of the previous 23 16 day intervals). | Aggregated into quartiles for multivariable regressions. |
| *NDVI* | This is the normalized difference vegetation index, detailed in the Supporting materials. | Mean values at 2km, 5km, and 10km buffers around each village. | For village level analysis: calculated as a mean value for the study duration. For individual level analysis: Calculated for the same month (same 16 day time period); the previous 2 months (mean of the previous 5 16 day intervals); and the previous year (mean of the previous 23 16 day intervals). |  |
| *EVI* | This is the enhanced vegetation index, detailed in the Supporting materials. | Mean values at 2km, 5km, and 10km buffers around each village. | For village level analysis: calculated as a mean value for the study duration. For individual level analysis: Calculated for the same month (same 16 day time period); the previous 2 months (mean of the previous 5 16 day intervals); and the previous year (mean of the previous 23 16 day intervals). | Aggregated into quartiles for multivariable regressions. |
| *Gender* | Binary for male or female, self reported in hospital records |  |  |  |
| *Age* | Self reported age in years. |  |  | Aggregated into age groups for multivariable |
| *Year* | The year of admission to the hospital |  |  | For multivariable regressions, this variable was centered on its mean and standardized by its standard deviation. |
| *Quarter* | The calendar quarter of admission (Jan - March; April - June; July - Sep; Oct - Dec) |  |  |  |

### Exploratory spatial data analysis

Summary statistics (median; Q1:Q3; mean) were calculated for the distances between villages and the nearest major road, and aggregated by whether or not the village was home to included patients and by specific diagnoses. Summary statistics (mean and 95% confidence intervals) were also calculated for all environmental variables, at each buffer size and temporal resolution, and for each of the major diagnoses. Tukey’s post hoc range test was used to assess statistically significant differences in environmental indices across diagnoses.

Standard distance deviations (SDDs) and standard deviation ellipses (SDEs) were calculated (**details in Supporting Materials II**) and mapped to measure and visually analyze the central tendency and spatial distributions for all patient home villages and by each of the major single (mono-infection) diagnoses.

### Formal analyses

Multivariable regressions were used to calculate model-adjusted odds ratios and confidence intervals. The regressions at the village level focused on study patient villages and the home villages of JEV diagnosed patients. A multivariable regression was also done at the individual level focusing on JEV infected patients. Other diagnoses were not included in these analyses because of small numbers of cases per village.

Logistic generalized additive models (GAMs) were used for variable selection and specification (**detailed in Supporting Materials IV and in Table 1**) for the final models. The GAMs were used to examine different specifications of the continuous environmental, geographic, and demographic variables and for changes in model fit and strength of association across buffer sizes (i.e. 2km, 5km, or 10km buffers) and for different time durations for EI measurements (i.e. same month, 2 months prior, 12 months prior to hospital admission). The final model covariates were chosen based on a combination of *a priori* hypotheses, model fit (using the Akaike information criterion), and strength of association between the covariate and the model outcome variable.

The final model for the individual-level analysis was a generalized logistic mixed model with a random effect for home village. The final model for the village-level analysis was a logistic regression.

### Software

All maps were created using QGIS version 3.4.9. R cran version 3.5.2 was used for downloading, processing, and wrangling MODIS data (using the “MODIStsp”; “raster”; “rgdal”; and “maptools” packages) and for all regressions. The “mgcv” package was used for GAMs and the lme4 package was used for the generalized mixed models. Euclidian distances between villages and major roads were calculated using QGIS. ArcMap version 10.5.1 was used to calculate SDDs and SDEs.

### Ethics approval

Ethical clearance for the CNS study was granted by the Oxford University Tropical Ethics Research Committee and by the Ethical Review Committee of the Faculty of Medical Sciences, National University of Laos. Between 2003 and 2006, both Oxford Tropical Ethics Committee and the Ethical Review Committee of the Faculty of Medical Sciences, National University of Laos, approved the use of oral witnessed consent. Oral consent was confirmed by a signed witness statement. Verbal consent (from 2003 – 2006) and written consent (from 2006 – 2011) were obtained from all recruited patients or immediate relatives. All ages were included provided that they or their guardian gave witnesses oral informed consent (2003 – 2006) and written informed consent since 2006.

## RESULTS

### Summary statistics

A total of 1,065 patients were recruited with no LP contraindications and consented to have a diagnostic LP; 450 (42%) were assigned a final laboratory diagnosis. The most common of these were Japanese encephalitis virus ((JEV) 94 individuals); followed by *Cryptococcus* spp. with 70 individuals (9 were *C. gattii*)); scrub typhus (*Orientia tsutsugamushi*) 31; Dengue virus 27; *Leptospira* spp. 25; murine typhus (*Rickettsia typhi*) 24; *Streptococcus pneumoniae* in 22 and 20 with *Mycobacterium tuberculosis*. 124 patients died prior to discharge (out of 893 with recorded discharge type recorded).

The majority (666, 63%) of patients were male, with the lowest sex bias in cryptococcal infections (40/70, 57% male) and the highest among dengue infections (22/27, 82% male) (**Table 2**). Age patterns were evident in JEV and cryptococcal infections, with median ages of 13 and 33 years, respectively (**Table 2**). Patients were linked to 582 different villages (multiple patients could come from the same village): 90 villages with JEV patients, 66 with cryptococcal patients, 31 with scrub typhus patients, 27 with dengue patients, 24 with leptospiral patients, and 24 with murine typhus patients. The majority (870, 82%) of patients came from within Vientiane Prefecture (678, 64%) or Vientiane Province (192, 18%).

**Table 2:** Age and gender of study patients. (Q1 and Q3 indicate the first and third quartiles, respectively).

|  | male/female | M/F ratio | median age in years (Q1 –Q3) | Total number |
| --- | --- | --- | --- | --- |
| **all patients** | 666/399 | 1.67 | 23 (8 - 38) | 1065 |
| **JEV** | 55/39 | 1.41 | 13 (8 - 20) | 94 |
| ***Cryptococcus* spp.** | 40/30 | 1.33 | 33 (27 - 41) | 70 |
| **scrub typhus** | 22/9 | 2.44 | 16 (9 - 29) | 31 |
| ***Dengue virus*** | 22/5 | 4.4 | 20 (9 - 30) | 27 |
| ***Leptospirosis* spp.** | 17/8 | 2.13 | 25 (12 - 39) | 25 |
| **murine typhus** | 17/7 | 2.43 | 32 (16 - 51) | 24 |

A total of 6,416 villages (of 10,520 recorded in 2005 [16]) were selected as the study area for further village level analyses (**Table 3**). Villages that were home to study patients were 11 times (0.7km versus 6.3km, from **Table 3**) closer to a major road when compared to other villages within the study area. Scrub typhus and JEV infected patient homes were further from major roads than other patients, but the difference was not statistically significant in univariate analyses.

**Table 3:** Distribution of distances (in km) to the nearest major road, by diagnosis type. Counts of villages are from within 3 standard deviational ellipses (SDEs) of all LP villages (referred to as the “study area” in text). In some cases, multiple patients came from the same village meaning that counts of villages will be smaller than counts of total patients. (Q1 and Q3 indicate the first and third quartiles, respectively).

|  | n | median distance in km (Q1 - Q3) |
| --- | --- | --- |
| **All** | 6416 | 5.4 (0.5 - 15.2) |
| **Villages without study patient** | 5847 | 6.3 (0.8 - 16.1) |
| **Villages with study patient** | 569 | 0.7 (0.1 - 4.1) |
| **JEV** | 88 | 0.6 (0.1 - 8.0) |
| ***Cryptococcus* spp.** | 66 | 0.3 (0.1 - 1.4) |
| **scrub typhus** | 31 | 0.6 (0.2 - 3.5) |
| ***Dengue virus*** | 27 | 0.3 (0.1 - 1.1) |
| ***Leptospirosis* spp.** | 22 | 0.4 (0.1 - 1.9) |
| **murine typhus** | 24 | 0.3 (0.1 - 2.2) |

The home villages of JEV patients were more broadly dispersed (**Figure 1B**) than for patients with other etiologies (**Figure 1C**), evident from the circular, larger SDE and SDD. The distribution of these JEV patient home villages was also relatively isotropic, with the SDE and SDD being nearly identical. Conversely, patients with cryptococcal infections were clustered near Vientiane City and along the road leading North/Northwest from the urban center (**Figure 1C**). Scrub typhus and murine typhus infections were also both clustered around Vientiane City but showed perpendicular spatial distributions (**Supporting Figures 4D and 4G**) a pattern previously described from IgG seropositivity data from Vientiane City [17]).

### Characteristics of patient home villages

Mean NFI values for the 2km radius tended to be higher than for either the 5km or 10km radius as surface flooding is heterogeneous and taking a mean across larger radii dilutes the measurement. The opposite pattern was observed for both vegetation indices. The 2km radius for both mean NDVI and mean EVI was usually smaller than at 5km or 10km radii (**Figures 2 and 3**).

Study patient villages had higher mean NFI values than non-study patient villages (**Figure 2A**) (non-study patient villages are those in the same study area as study patient villages but were not home to a study patient). The home villages of study patients diagnosed with *dengue virus* and cryptococcal infections had high mean NFI values over the duration of the study period when compared to other major diagnoses (**Figure 2A**).

Conversely, the home villages of study patients tended to have lower mean EVI values than non-study patient villages (**Figure 2C**). Villages from which patients who were diagnosed with JEV were an exception to this general pattern. JEV patient home villages had higher mean EVI when compared to home villages of patients with dengue and cryptococcal infections (**Figure 2C**).

Home villages of patients with JEV diagnoses had lower mean NFI over the duration of the study period, but had higher NFI than other major diagnoses when looking specifically at the month of admission (especially when compared to cryptococcal infections and murine typhus (**Figure 3A**)). JEV patient home villages also had higher EVI during the month of admission than most other mono-infections, especially when compared to either cryptococcal infections or murine typhus (**Figure 3C**). Scrub typhus infections had higher EVI than cryptococcal infections when the measurement was taken at the 10km radius buffer (not detectable at smaller radii (**Figure 3C**)).

Home villages of patients with dengue infections had higher NFI than murine typhus or *Cryptococcus* spp. patient home villages when considering the 2 months prior to admission (**Figure 3D**). *Cryptococcus* spp. patient home villages had particularly low EVI in the two months leading up to admission, especially when compared to JEV and *Leptospira* spp. patient home villages (**Figure 3F**). At one year prior to admission both *Cryptococcus* spp. patient home and *dengue virus* patient home villages had higher NFI than JEV patient home villages (**Figure 3G**).

### Logistic regressions for geographic, environmental and demographic predictors of CNS infections

In agreement with univariate analyses, villages from which study patients came tended to be larger in population size (**Supporting Figure 4**), lower in elevation (**Supporting Figure 5**), and closer to a major road when compared to other villages within the study area (**Table 4**). They also had higher mean levels of surface flooding, with villages in the highest NFI quadrant having over two times the odds (AOR: 2.21; CI: 1.49 – 3.31) of being a home village for study patients when compared to neighboring villages in the study area (**Table 4 and Figure 2**). Villages from which JEV patients originated had few defining characteristics in the logistic regression, other than being larger in population size (AOR: 1.74; CI: 1.55 – 1.96) and at lower elevations (AOR: 0.69; CI: 0.46 – 0.97) than non-study patient villages (**Table 5**).

**Table 4:** Logistic regression and model adjusted odds ratios (AOR) for village level analysis of LP villages

| covariate | total | LP count | AOR (CI) |
| --- | --- | --- | --- |
| NDFI Q1 | 1604 | 68 | reference group |
| NDFI Q2 | 1604 | 75 | 1.07 (0.73 - 1.56) |
| NDFI Q3 | 1604 | 130 | **1.48 (1.03 - 2.16)** |
| NDFI Q4 | 1604 | 296 | **2.21 (1.49 - 3.31)** |
| EVI Q1 | 1604 | 311 | reference group |
| EVI Q2 | 1604 | 134 | 1.16 (0.86 - 1.57) |
| EVI Q3 | 1604 | 74 | 1.12 (0.76 - 1.66) |
| EVI Q4 | 1604 | 50 | 1.19 (0.74 - 1.91) |
| Village population |  |  | **2.22 (2.03 - 2.42)** |
| Elevation |  |  | **0.52 (0.42 - 0.63)** |
| Distance to major road |  |  | **0.68 (0.57 - 0.80)** |

**Table 5:** Logistic regression and model adjusted odds ratios (AOR) for village level analysis of JEV villages

| covariate | total | JEV count | AOR (CI) |
| --- | --- | --- | --- |
| NDFI Q1 | 1604 | 18 | reference group |
| NDFI Q2 | 1604 | 15 | 0.83 (0.38 - 1.76) |
| NDFI Q3 | 1604 | 20 | 1.11 (0.53 - 2.35) |
| NDFI Q4 | 1604 | 35 | 1.26 (0.54 - 2.95) |
| EVI Q1 | 1604 | 35 | reference group |
| EVI Q2 | 1604 | 23 | 1.81 (0.87 - 3.76) |
| EVI Q3 | 1604 | 17 | 1.85 (0.77 - 4.46) |
| EVI Q4 | 1604 | 13 | 1.76 (0.63 - 4.93) |
| Village population |  |  | **1.74 (1.55 - 1.96)** |
| Elevation |  |  | **0.69 (0.46 - 0.97)** |
| Distance to major road |  |  | 0.88 (0.62 - 1.20) |

In the individual-level analysis (**Table 6**), age and season were the strongest predictors of JEV infections among all patients. Patients who were admitted between July and September had over seven times the odds (AOR: 7.40; CI: 1.45 – 37.67) of being diagnosed with JEV when compared to patients who were admitted between January and March (**Table 6**). JEV was most common in children aged 5 through 14 (AOR: 2.74; CI: 1.31 – 5.69; ages 0 – 4 as the comparison group). NFI during the month of admission (10km buffer used in the regression) was also a strong predictor of JEV infection. Individuals who came from villages in the highest quadrant of NFI measurements had approximately 3 times the odds being diagnosed with JEV (AOR: 3.06; CI: 1.04 – 8.96). While study patients came from villages with lower mean elevations, patients who were diagnosed with JEV came from higher elevation villages in comparison to the other patients (AOR: 1.36; CI: 1.11 – 1.66). EVI was a significant predictor in models that did not include distance to road, village population, and elevation (**Table 6 M1 and M2**).

**Table 6:** Mixed effects logistic regression and model adjusted odds ratios (AOR) for individual level analysis

|  |  |  | **M1** | **M2** | **M3** |
| --- | --- | --- | --- | --- | --- |
| **covariate** | **total** | **JEV count** | **AOR (CI)** | **AOR (CI)** | **AOR (CI)** |
| NDFI Q1 | 262 | 7 | reference group | reference group | reference group |
| NDFI Q2 | 261 | 16 | **2.91 (1.06 - 7.97)** | 2.28 (0.86 - 6.01) | 2.32 (0.87 - 6.19) |
| NDFI Q3 | 261 | 31 | **2.73 (1.02 - 7.28)** | **2.75 (1.07 - 7.07)** | **2.98 (1.13 - 7.85)** |
| NDFI Q4 | 262 | 38 | **3.41 (1.17 - 9.94)** | **3.12 (1.09 - 8.90)** | **3.06 (1.04 - 8.96)** |
| EVI Q1 | 262 | 8 | reference group | reference group | reference group |
| EVI Q2 | 261 | 20 | 2.11 (0.84 - 5.28) | 1.89 (0.76 - 4.74) | 1.60 (0.63 - 4.04) |
| EVI Q3 | 261 | 24 | 2.04 (0.81 - 5.12) | 1.65 (0.67 - 4.07) | 1.33 (0.53 - 3.32) |
| EVI Q4 | 262 | 40 | **4.19 (1.62 - 10.87)** | **3.44 (1.35 - 8.73)** | 2.43 (0.91 - 6.46) |
| Jan - March | 210 | 2 | reference group | reference group | reference group |
| April - June | 267 | 22 | **5.12 (1.10 - 23.89)** | 4.45 (0.95 - 20.76) | **5.05 (1.07 - 23.73)** |
| July - Sep | 333 | 62 | **8.72 (1.75 - 43.46)** | **6.35 (1.26 - 31.89)** | **7.40 (1.45 - 37.67)** |
| Oct - Dec | 253 | 8 | 1.80 (0.35 - 9.25) | 1.51 (0.29 - 7.81) | 1.54 (0.29 - 8.17) |
| Year |  |  | **1.31 (1.03 - 1.68)** | **1.34 (1.05 - 1.72)** | 1.27 (0.98 - 1.64) |
| 0 through 4 | 208 | 13 |  | reference group | reference group |
| 5 through 14 | 150 | 37 |  | **3.00 (1.46 - 6.18)** | **2.74 (1.31 - 5.69)** |
| 15 through 24 | 192 | 28 |  | 1.74 (0.84 - 3.62) | 1.37 (0.64 - 2.94) |
| 25 through 34 | 186 | 10 |  | 0.66 (0.27 - 1.61) | 0.61 (0.25 - 1.48) |
| 35 through 44 | 133 | 4 |  | **0.26 (0.07 - 0.96)** | **0.26 (0.07 - 0.97)** |
| 45 plus | 196 | 2 |  | **0.11 (0.02 - 0.52)** | **0.10 (0.02 - 0.47)** |
| female | 399 | 39 |  | reference group | reference group |
| male | 666 | 55 |  | 0.97 (0.60 - 1.58) | 1.08 (0.66 - 1.79) |
| Village population |  |  |  |  | 1.00 (0.77 - 1.29) |
| Elevation |  |  |  |  | **1.36 (1.11 - 1.66)** |
| Distance to major road |  |  |  |  | 1.08 (0.88 - 1.32) |

## DISCUSSION

Patients from this study were recruited based on symptomology and a medical procedure that is only available at a single location in the nation (diagnostic LP at Mahosot Hospital, Vientiane). The home villages of all included study patients, regardless of diagnosis, were approximately centered on Vientiane City and were closer to major roads than would be expected by chance alone. For many of the infections studied in this analysis, this association is likely more related to geographic and financial access to healthcare systems rather than exposure to environmental risk factors – especially for infections that are more associated with rural areas (e.g. JEV). This finding also suggests that the results here may be a vast underestimate of the true burden of CNS infections, with much of the Lao population not being in near proximity to a major road (**Table 3**). The causative agents of CNS infections differ in biology, ecology, and geography, and this is evident through the spatial distributions of the home villages of patients. The geographic, environmental, and demographic patterns exhibited by patients needing a diagnostic LP for suspected CNS infections, and for specific diagnoses, are the result of complex overlapping factors.

A similar spatial pattern was described from an epidemiological analysis of CNS infections among children admitted to Ho Chi Minh City hospitals in Vietnam – with most patients coming from districts near the hospital [18]. While the majority of infections (55%) in the Vietnam study were presumed to be bacterial in origin, in this study from Lao PDR bacterial infections were identified in only 38% (170 out of 450 patients with diagnoses).

JEV was the single largest identified cause of CNS infections in these data; it primarily affected children (median 13 years of age, **Table 2**), occurred predominantly during the rainy season (likely corresponding to peaks in mosquito vector populations), and in villages with recent high levels of surface water [19]. JEV is a vaccine-preventable disease, but the vaccine has historically been expensive and vaccine programs are frequently limited by access to remote communities. In 2013 the WHO approved a less expensive vaccine (produced in China by the Chengdu Institute of Biological Products) which has since been used in mass vaccination campaigns in Lao PDR in 2013 and 2015 [20]. The vaccine is now routinely given to all children less than 9 months of age but coverage may be low in some areas.

The second largest contributor to CNS diseases were cryptococcal infections, which are opportunistic fungal infections with high mortality [21]. Of the 70 patients with cryptococcal infections, 12 died prior to discharge and another 8 likely died at home after leaving the hospital. Cryptococcal infections are generally acquired after inhalation of the yeast-like form of the fungus which has been associated with several ecological habitats (*Cryptococcus gattii* has been associated with over 50 species of trees; *Cryptococcus neoformans* has been associated with bird droppings but is also suspected to be associated with plants [22,23]). This disease has a long incubation period [24,25] and while *C. gattii* infections commonly occur among immunocompetent individuals, *C. neoformans* infections are frequently associated with HIV infections [26,27]. In these data, 20 of the patients diagnosed with cryptococcal infections also had HIV infections. While several studies have shown that cryptococcal species exist in specific ecological habitats and have inferred environmental exposure, the long incubation period and complex natural history likely obfuscate ecological correlations.

There are several limitations to this research. Diagnostic LPs are only conducted in Mahosot Hospital in the national capital. Logistical and financial difficulties in accessing healthcare facilities, and especially for etiological diagnostic capabilities, likely leads to severe under-reporting of meningitis, encephalitis, or in the diagnosing the causative agent in these conditions when the patient does access care. All of these factors ultimately lead to small case counts for numerous different causative agents. The spatial patterns in points (villages) and ellipses exhibited in these data are likely influenced by the shape of the nation and it is possible that the point patterns and ellipses would differ if we had data from neighboring nations. Spatial and temporal patterns that differentiate different infections might be more obvious if the surveillance system instead focused on any symptomatic infections (rather than only suspected infections of the CNS). Some pathogens are neurotropic whereas others have tropism for other organs, while being capable of occasionally infecting the CNS. This may partially explain the higher case counts of JEV and why we were able to identify spatial, temporal, and environmental predictors for this causative agent.

OpenStreetMaps data are volunteered data and may be prone to error. For this reason we focused on major roads, whose routes have changed very little over the last decades. For the regressions, the distances from all villages to the nearest major road was also rounded to the nearest 5km. Examination of satellite imagery in comparison with the major roads from OpenStreetMaps suggests that where error does exist, it is on a scale of ±100 meters, meaning that measurements of distances, as used in this analysis, should not be strongly influenced. Some of these data now come from over a decade ago. Surveillance systems of this type (based on relatively vague symptomology), with a wide panel of possible contributing causative agents, and necessary intensive laboratory components are extremely labor and time intensive.

Lao PDR is currently undergoing vast environmental, demographic, and economic changes. Road networks are increasing in range and density and several areas (i.e. Vientiane, Savannakhet) are undergoing expansive urbanization [28]. These environmental changes will most likely result in shifting patterns of infectious diseases. As the region undergoes urbanization (including both a decrease in urban landscape and movement of human populations to urban centers), pathogens that thrive in rural areas (e.g. JEV) may undergo reduced transmission, especially if vaccine campaigns are more capable of reaching rural populations. Conversely, infections that cluster in urban and peri-urban areas (such as dengue and murine typhus) may increase in frequency.

Several environmental indices from remote sensing instruments have shown potential for predicting disease risk, differentiating disease types, or for other surveillance efforts in SE Asia [29,30] and globally [31–35]. This analysis, and others like it, illustrates the ability to differentiate some infections (namely JEV when compared to other diagnoses) through the use of freely available data (i.e. MODIS) and software (R and QGIS) and routinely collected healthcare data. Surveillance systems and potentially diagnostic algorithms [36] in developing settings could benefit from inclusion of such resources. A far-reaching surveillance system that is representative of the entire nation and includes likely CNS infections would be beneficial in order to assess the true burden of CNS infections – many of which would benefit from primary and secondary prevention through increased provision of vaccines, vector control, and early diagnosis and treatment. Given the inherent difficulties in accurately diagnosing and treating CNS infections, the predictors reported here and from other epidemiological studies for major contributors to CNS diseases (i.e. age, seasonality, location, and environmental characteristics) could be considered alongside clinical symptomology when presumptive diagnoses are being made. However, it will be important to consider current and ongoing demographic, environmental, and economic changes in Lao PDR.

Finally, increasing population access to vaccines, diagnosis, and treatment would have clear benefits to overall population health. As with other parts of the developing world, a large fraction of the Lao population must travel long distances in order to reach primary healthcare centers. In 2005 73% of the Lao population was reported to live in rural areas, 21% without roads. By 2015 67% of the population were reported to live in rural villages with 8% in villages without roads [37]. For many communities, travel during the wet season remains difficult. Travel costs can also be prohibitive. Most of the CNS infections in this analysis occurred or developed symptoms during the wet season. Public health initiatives that help to decrease the distances between communities and the healthcare services that they need are warranted.

## Data Availability

Data from this manuscript come from four different sources.
1.) Lao Bureau of Statistics Census data, available through: http://www.decide.la/en/
2.) Remote sensing data available through: https://modis.gsfc.nasa.gov/data/
3.) Roads and highway data available through: http://www.openstreetmap.la
4.) Patient hospital records which can be requested from the Mahidol Oxford Tropical Medicine Research Unit (following policies and guidelines): http://www.tropmedres.ac/data-sharing

https://modis.gsfc.nasa.gov/data/

http://www.openstreetmap.la

http://www.tropmedres.ac/data-sharing

## Acknowledgements

We are very grateful to the patients and to Bounthaphany Bounxouei, past Director of Mahosot Hospital; to Bounnack Saysanasongkham, Director of Department of Health Care, Ministry of Health; to H.E. Bounkong Syhavong, Minister of Health, Lao PDR; to the staff of the Infectious Disease Center and Microbiology Laboratory for their help and support.

## Supporting information legends

**Supporting information file 1: Detailed explanation of variables and analyses**

## FIGURES

**Figure 1:** Spatial distributions of the home villages of study patients, for A: all study patients, B: study patients with JEV infections, and C: with cryptococcal infections, D: scrub typhus infections, E: with *dengue virus* infections, F: with leptospiral infections, and G: with murine typhus infections. SDDs and SDEs are weighted by case numbers, with some patients coming from the same village. Maps were created using QGIS version 3.4.9. All layers were created by the authors of this manuscript.

**Figure 2:** Environmental indices for villages with study patient homes for the duration of the study period (January 2003 through August 2011) for all study patient villages, non study patient villages in the study area, and for major diagnoses (JEV = Japanese Encephalitis virus; Crypto = cryptococcal infection; ST = scrub typhus; MT = murine typhus; dengue = Dengue virus; lepto = *Leptospira* spp. infection). Bar values are mean values and the error bars are 95% confidence intervals, using the t-distribution. NFI values here have a constant (0.25) added to them for visualization only.

**Figure 3:** Environmental indices for study patients by major diagnosis and at different times leading up to the date of admission. JEV = Japanese Encephalitis virus; Crypto = cryptococcal infection; ST = scrub typhus; MT = murine typhus; dengue = *Dengue virus*; lepto = *Leptospira* spp. infection. Bar values are mean values and the error bars are 95% confidence intervals, using the t-distribution. NFI values here have a constant (0.25) added to them for visualization only.

